# A practical guideline of genomics-driven drug discovery in the era of global biobank meta-analysis

**DOI:** 10.1101/2021.12.03.21267280

**Authors:** Shinichi Namba, Takahiro Konuma, Kuan-Han Wu, Wei Zhou, Global Biobank Meta-analysis Initiative, Yukinori Okada

## Abstract

Genomics-driven drug discovery is indispensable for accelerating the development of novel therapeutic targets. However, the drug discovery framework based on evidence from genome-wide association studies (GWAS) has not been established, especially for cross-population GWAS meta-analysis. Here, we introduce a practical guideline for genomics-driven drug discovery for cross-population meta-analysis, as lessons from the Global Biobank Meta-analysis Initiative (GBMI). Our drug discovery framework encompassed three methodologies and was applied to the 13 common diseases targeted by GBMI (*N*_mean_ = 1,329,242). First, we evaluated the overlap enrichment between disease risk genes and the drug-target genes of the disease-relevant medication categories. An omnibus approach integrating the four gene prioritization tools yielded twice the enrichment in the disease-relevant medication categories compared with any single tool, and identified drugs with approved indications for asthma, gout, and venous thromboembolism. Second, we performed an endophenotype Mendelian randomization analysis using protein quantitative trait loci as instrumental variables. After the application of quality controls, including a colocalization analysis, significant causal relationships were estimated for 18 protein–disease pairs, including MAP2K inhibitors for heart failure. Third, we conducted an *in silico* screening for negative correlations between genetically determined disease case–control gene expression profiles and compound-regulated ones. Significant negative correlations were observed for 31 compound–disease pairs, including a histone deacetylase inhibitor for asthma. Integration of the three methodologies provided a comprehensive catalog of candidate drugs for repositioning, nominating promising drug candidates targeting the genes involved in the coagulation process for venous thromboembolism. Our study highlighted key factors for successful genomics-driven drug discovery using cross-population meta-analysis.

## Introduction

Efficient screening of novel therapeutic targets is an essential process to accelerate drug discovery. Despite the enormous effort to develop novel drugs, the overall success rate of clinical application has been decreasing because of the considerable increase in both the cost and the duration^1^. Genomics-driven drug discovery is one of the promising solutions, as drug targets with human genetic support are more likely to be successful in clinical development^2,3^. In particular, rare-variant studies for mendelian diseases have led to drug development, such as *PSCK9* inhibitors for low-density lipoprotein cholesterol^4^. For common diseases, genome-wide association studies (GWAS) have provided valuable opportunities for drug discovery; nevertheless, drug discovery based on GWAS remains challenging^5^. Few bioinformatics tools directly prioritize candidate drugs^6^, and there exist no practical guidelines regarding how to conduct genomics-driven drug discovery.

Recently, an increasing number of large-scale GWAS meta-analyses of multiple populations have been carried out. These have revealed key insights into the biological processes underlying complex diseases^7,8^, thus affording the possibility of in-depth application of genomics-driven drug discovery. However, the majority of previous genomics-driven drug discovery projects were carried out for GWAS of a single ancestry of Europeans^6^, and there are few successful applications to cross-population GWAS meta-analyses. The global heterogeneity in genetic background (e.g., different allele frequencies and linkage disequilibrium [LD]) among populations makes it difficult to perform downstream analyses such as gene expression prediction^9^ and colocalization analysis^10^. In addition, causal effect sizes are population-specific especially in functionally important regions^11^. Therefore, a specialized drug discovery framework is required for cross-population GWAS meta-analyses. In this study, we introduce a practical guideline as lessons from the Global Biobank Meta-analysis Initiative (GBMI)^7^. GBMI meta-analyzed GWASs of global biobanks from diverse ancestries incorporating several recruitment strategies (e.g., population-based or hospital-based biobanks), including up to 1.8 million participants from the 18 biobanks in four continents (341,000 East Asians (EAS); 31,000 Central and South Asians; 33,000 Africans; 18,000 Admixed Americans; 1,600 Middle Easterners; 156,000 Finns; and 1,220,000 Non-Finnish Europeans [NFE]), serving as a gold-standard cross-population GWAS meta-analysis. We propose a cross-population drug discovery framework comprising three major methodologies. First, overlap enrichment of disease risk genes with targets of existing drugs^12–14^ identifies drug repurposing opportunities. Second, endophenotype Mendelian randomization (MR), and subsequent quality controls, including colocalization analyses^15^ establishes causal links between proteins and disease processes. Lastly, screening of negative correlations between genetically regulated disease case–control gene expression (GReX) and compound-regulated gene expression profiles^16^ can be used to identify compounds that might correct disease-related alterations in gene expression.

We applied our framework to the 13 common and relatively rare disease GWAS included in GBMI: asthma, primary open-angle glaucoma (POAG), gout, chronic obstructive pulmonary disease (COPD), venous thromboembolism (VTE), thyroid cancer (ThC), abdominal aortic aneurysm (AAA), heart failure (HF), idiopathic pulmonary fibrosis (IPF), stroke, uterine cancer (UtC), acute appendicitis (AcApp), and hypertrophic cardiomyopathy (HCM). Our pipeline identified 144 drug/compound-disease pairs and nominated a comprehensive catalog of candidate drugs for repositioning. Our study demonstrates that the integration of the candidate drugs and compounds screened using each methodology enables in-depth drug discovery. In particular, drug discovery captured plausible drug targets targeting coagulation-related genes for VTE. Our results demonstrated the utility of genomics-driven drug discovery using a cross-population GWAS meta-analysis and suggest key factors for successful drug discovery.

## Results

### Overview of the genomics-driven drug discovery framework

Various types of omics-based approaches have been proposed for novel target identification and drug repositioning from GWAS summary statistics^6^. Each type of omics data necessitates specialized methodologies. Further, no simple method has emerged that enables the interpretation of genomics-discoveries for drug discovery. Therefore, our framework was composed of three parts, in which each component utilized different external omics data and knowledge bases to obtain biological insights from GWAS summary statistics (**Figure 1**), as follows. (i) First, we performed an overlap enrichment analysis of the disease risk genes with the target genes of existing drugs^12–14^. Gene prioritization tools summarized variant-level *P*-values at the gene level, and the pharmacological agents targeting the prioritized genes served as drug candidates. Genes that are modulated by approved drugs are known to be enriched in disease-relevant clinical classifications, such as drug medication categories (i.e., Anatomical Therapeutic Chemical Classification System [ATC]) and disease categories (i.e., International Statistical Classification of Diseases and Related Health Problems [ICD-10])^13,14^, which were utilized to assess the validity of the drug candidates. (ii) Second, we performed endophenotype MR and subsequent quality controls, including colocalization analyses^15^. MR is a method that is used for estimating the causal effect of one trait (exposure) on another trait (outcome) using genetic variants as instrument variables (IVs)^17^. We used the lead variants of the protein quantitative trait loci (pQTL) as IVs to examine the disease-causing effects of the proteins. Subsequent quality controls were effective in avoiding false-positive causalities^18^. Particularly, colocalization analysis is an important step for the exclusion of confounding by LD. (iii) Finally, we performed a screening of negative correlations between genetically regulated disease case–control gene expression (GReX) and compound-regulated gene expression profiles^16^. Transcriptome-wide association studies (TWAS) use expression QTL (eQTL) acting in *cis* to impute disease-specific GReX from the GWAS summary statistics^9^. The compounds that have inverse effects on gene expression against case–control GReX serve as candidates for the disease of interest^16,19^. We imputed GReX for the tissues included in the Genotype-Tissue Expression project (GTEx) v7^20^ and used compound-induced gene expression profiles for thousands of compounds in various conditions and cell lines collected in one of the largest public databases available, the Library of Integrated Network-based Cellular Signatures project (LINCS) L1000 connectivity map^21^.

**Figure 1.**
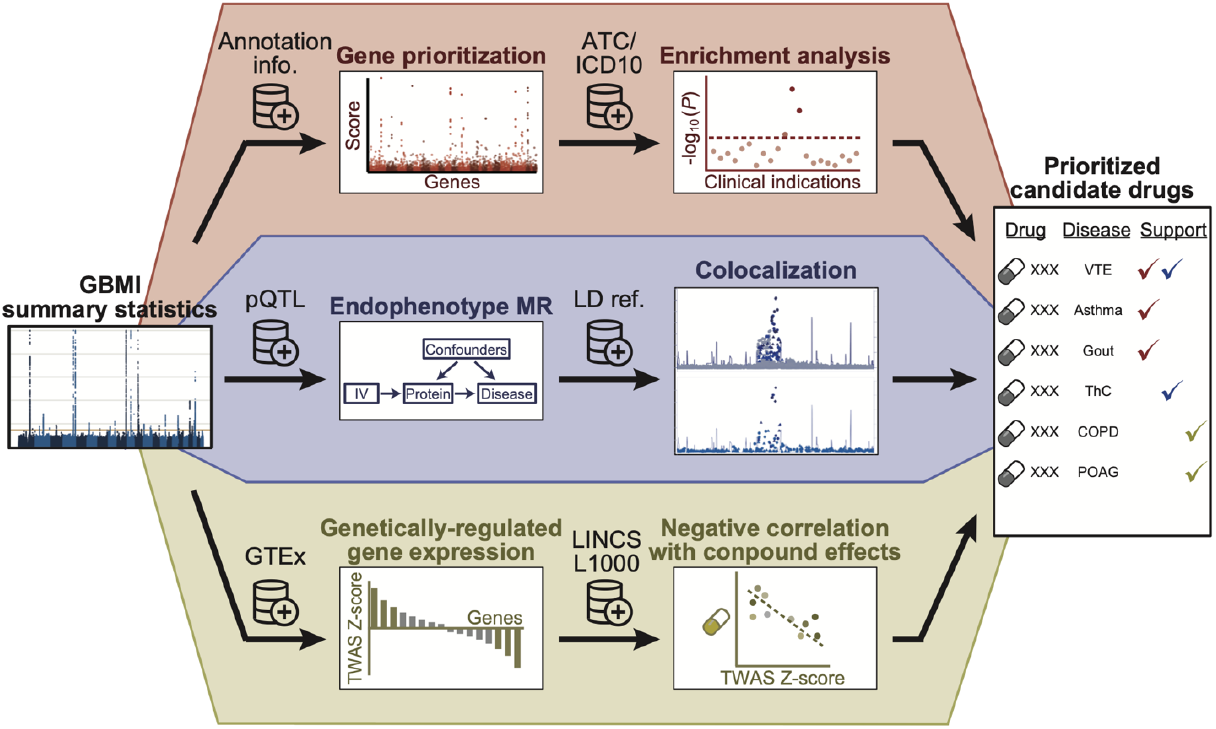
Overview of the genomics-driven drug discovery framework. The framework consisted of three components. Each component utilizes the summary statistics of genome-wide association analyses and external resources to prioritize candidate drugs. GBMI, Global Biobank Meta-analysis Initiative; ATC, Anatomical Therapeutic Chemical Classification System; ICD10, International Statistical Classification of Diseases and Related Health Problems; pQTL, protein quantitative trait loci; LD, linkage disequilibrium; MR, Mendelian randomization; GTEx, the Genotype-Tissue Expression project; TWAS, transcriptome-wide association study; LINCS, the Library of Integrated Network-based Cellular Signatures project.

### Overlap enrichment of disease risk genes in medication categories

The first component prioritized disease risk genes by calculating gene scores or *P*-values, and examined whether the prioritized genes are enriched in drug-target genes of specific medication categories. There exist several gene prioritization tools, although it remains unclear which tool is best optimized for drug discovery. Therefore, we evaluated four tools in parallel, i.e., MAGMA^22^, DEPICT^23^, Priority index (Pi)^5^, and Polygenic Priority Score (PoPS)^24^. MAGMA is a simple method that is used to summarize variant-level *P*-values according to gene positions and LD structure. The prioritized genes by MAGMA were nominally overlapped with drug-target genes in the disease-relevant ATC codes for Gout, COPD, and VTE (*P* < 0.05; **Supplementary Figure 1A**). DEPICT, which uses co-regulated gene expression for gene prioritization^23^, showed clearer enrichment than did MAGMA for gout and VTE; however, no genes were prioritized for diseases with a relatively small number of genome-wide significant loci, such as IPF and UtC (**Supplementary Figure 1B**). Pi is a scoring system that was designed for drug development of immune-related diseases, and integrates multiple annotations, including eQTL, chromatin interaction, and genes implicated in immune functions^5^. The genes targeted by antineoplastic and immunomodulating agents were enriched for all diseases using Pi (**Supplementary Figure 1C**), suggesting that Pi could specifically provide enrichment of the immune genes regardless of the disease categories, even including non-immune diseases. PoPS estimates responsible genes using various gene features, such as cell-type-specific gene expression and biological pathways^24^. We observed enrichment of drug-target genes for broad ATC codes, suggesting that PoPS provided relatively high scores for the entire drug-target gene collection, regardless of their medication categories (**Supplementary Figure 1D**). We replicated the analyses using ICD-10 and observed a similar pattern of enrichment as ATC codes (**Supplementary Figure 2**).

Next, we summarized the overlap enrichment into disease-relevant and disease-irrelevant medication categories across the diseases. For both ATC and ICD-10 codes, all tools confirmed enrichment in the relevant codes, although a relatively high enrichment in the irrelevant codes was also observed for Pi and PoPS (**Figure 2A–C and Supplementary Figure 3**), reflecting biased enrichment in immune genes and non-specific enrichment in drug-target genes, respectively. DEPICT yielded a lower enrichment in irrelevant codes than did MAGMA. As a sensitivity analysis, we sequentially changed the thresholds of the gene scores and *P*-values. The pattern of the overall enrichment was robust for a wide range of thresholds (**Supplementary Figure 4**). The enrichment of disease genes prioritized by DEPICT in the relevant ATC codes became smaller with the stringent thresholds. DEPICT calculates *P*-values for the genes in the genome-wide significant loci exclusively, by default; therefore, liberal thresholds such as a false discovery rate (FDR) of 0.2 might be suitable for DEPICT.

**Figure 2.**
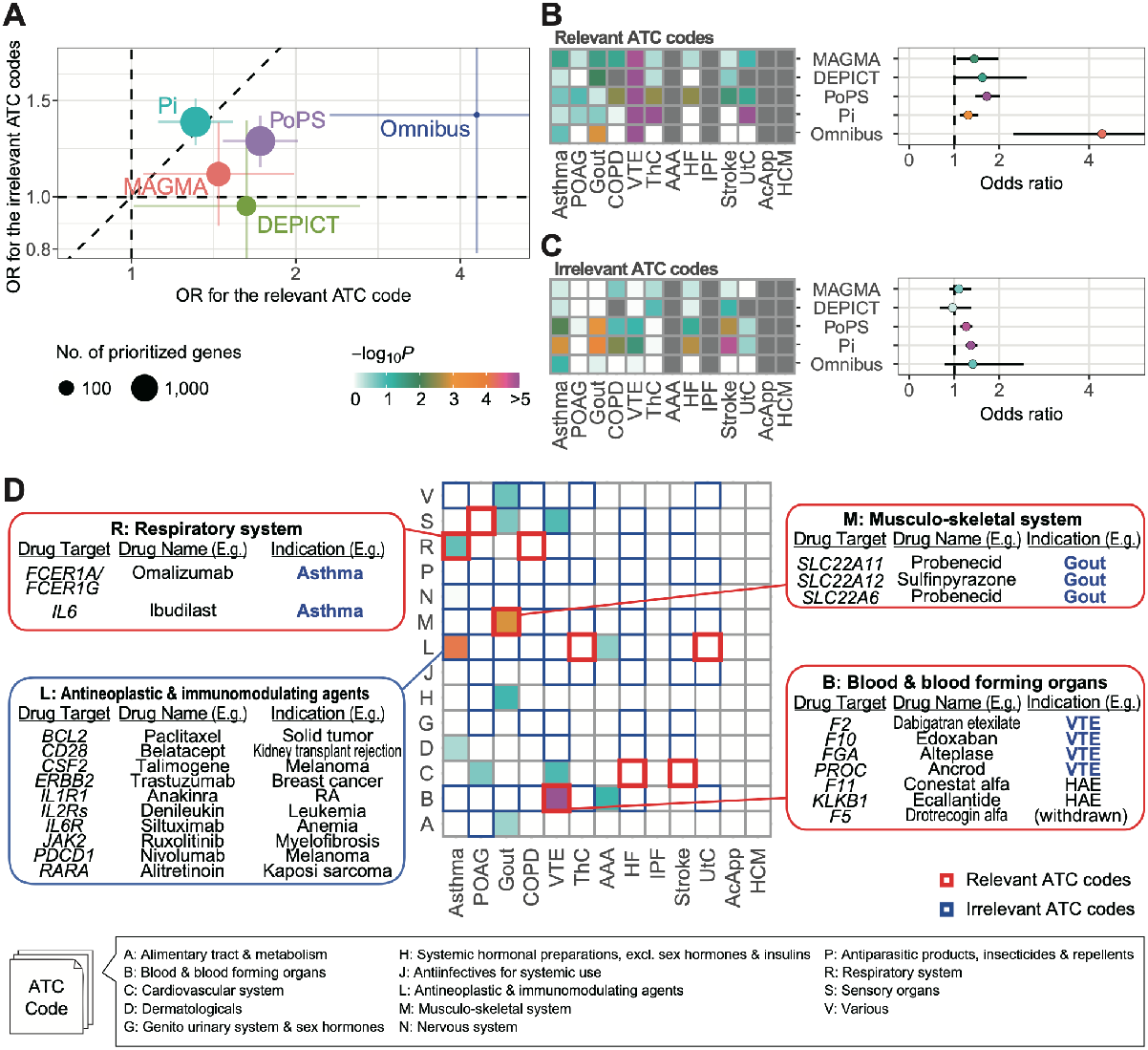
Enrichment of prioritized drug-target genes in the disease-relevant medication categories. **A**, Overall enrichment of drug-target genes nominated by four gene prioritization tools and their omnibus results. The error bars represent 95% confidence intervals. **B and C**, Enrichment of the prioritized drug-target genes in the disease-relevant ATC codes (**B**) and the disease-irrelevant ATC codes (**C**). The diseases are sorted in the descending order of the number of genome-wide significant loci determined in GBMI GWAS. **D**, Enrichments for the omnibus results per disease and ATC code. OR, odds ratio; POAG, primary open-angle glaucoma; COPD, chronic obstructive pulmonary disease; VTE, venous thromboembolism; ThC, thyroid cancer; AAA, abdominal aortic aneurysm; HF, heart failure; IPF, idiopathic pulmonary fibrosis; UtC, uterine cancer; AcApp, acute appendicitis; HCM, hypertrophic cardiomyopathy; RA, rheumatoid arthritis; HAE, acute attacks of hereditary angioedema.

Given that the four tools separately prioritized genes according to the different methodologies, we hypothesized that omnibus integration of the four methods could efficiently improve the enrichment of disease-relevant drug-target genes. As an omnibus approach, we selected 81 gene–disease pairs that were prioritized by all tools (**Supplementary Table 1**), and showed a twice larger OR than did those of any single tool for the relevant ATC codes (OR = 4.29, *P* = 5.5 × 10^−5^) (**Supplementary Figure 2A–C**). Conversely, the enrichment in the irrelevant ATC codes (OR = 1.41, *P* = 0.16) was close to that of the single tools, indicating an advantage of combining multiple gene prioritization methods to increase statistical power while retaining controlled type 1 errors. We also confirmed the efficacy of the omnibus strategy using ICD-10 (**Supplementary Figure 3**).

The examination of the omnibus results for each disease revealed an enrichment in the relevant ATC codes for asthma, gout, and VTE (**Figure 2D**), which corresponded to 30 drugs in total (**Supplementary Table 1**). For asthma and gout, all prioritized genes in the disease-relevant ATC codes were targeted by the drugs with approved indication. The genes prioritized for VTE were involved in the coagulation cascade, and four of them (*F2, F10, FGA*, and *PROC*) were the approved drug targets for VTE. The prioritized genes for asthma were also enriched in antineoplastic and immunomodulating agents. Although the drugs in this category have not been indicated for asthma, their target genes involved immune genes, such as *IL1R1*, suggesting that the omnibus approach correctly prioritized genes related to asthma.

### Endophenotype Mendelian randomization

Understanding protein regulation is essential for drug discovery. Therefore, we conducted MR analyses to infer disease-causing proteins^15,25^. Although the current eQTL studies yield larger sample sizes than pQTL^26^, there exists low correlation between transcript expression and protein abundance^27^. Here, we utilized the lead variants summarized from five pQTL studies^27–31^ as IVs for MR. To avoid confounding factors, such as horizontal pleiotropy, we restricted IVs to those with low heterogeneity between studies and low pleiotropy between proteins, whereas we used both *cis-* and *trans-*pQTL to target a wide range of proteins, as described previously^18^ (Methods). We conducted an MR analysis using the 266 IVs associated with the 229 drug-targeted proteins, to test the causal effects on the 13 diseases. After applying multiple test corrections, 25 protein–disease pairs showed significant causality (FDR < 0.05, **Supplementary Table 2**). Twelve pairs were derived from *cis*-pQTL, whereas the remaining ones were derived from *trans*-pQTL.

To protect against false positives from the results of the MR analysis, we applied two quality-control metrics, i.e., colocalization analysis and concordance of directional effects. The colocalization analysis was used to check whether two signals were equally distributed on the local LD structure. Therefore, we used the GBMI GWAS of the NFE-specific meta-analysis for MR and colocalization analysis, rather than the all-ancestry meta-analysis, to match the population background to the pQTL studies. All 25 pairs passed the directionality check, while colocalization was confirmed for 18 pairs (**Figure 3A**). These pairs included F11 and PROC for VTE and PLAU for AAA, which were also prioritized by the omnibus gene prioritization. In particular, F11 and PROC for VTE were also nominated by the overlap enrichment analysis. The colocalization of F11–VTE is shown in **Figure 3B** as an illustrative example. Most of the colocalization methods assume only a single causal variant per locus; however, recent methodological advances enabled us to address the possibility of multiple causal variants in one locus^32^. We applied coloc^33^ to conditionally independent signals decomposed by SuSiE^34^. SuSiE detected three and two signals for the GWAS of VTE and pQTL of F11, respectively, of which two signals were inferred to be colocalized.

**Figure 3.**
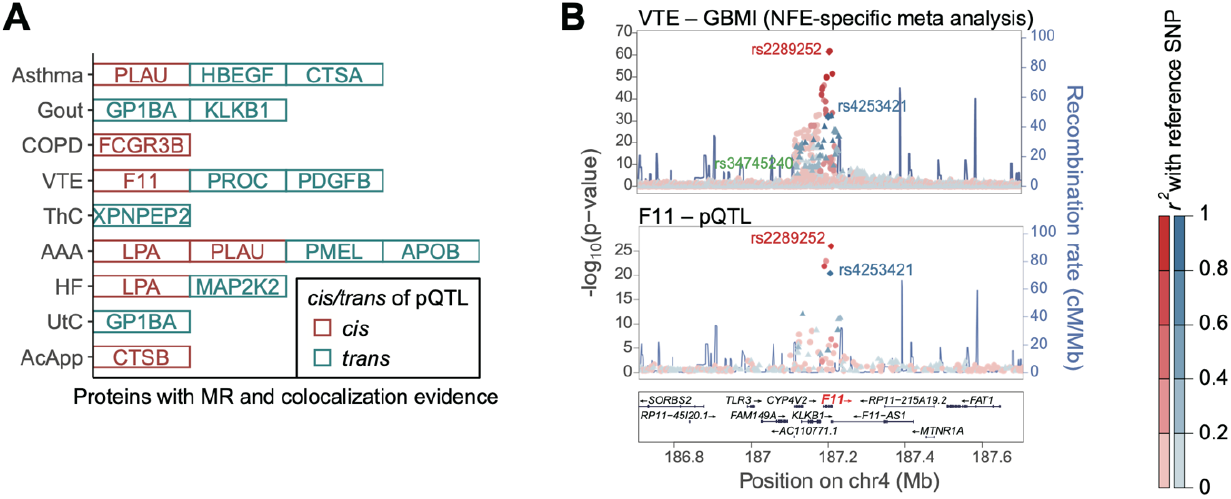
Endophenotype Mendelian randomization. **A**, Drug-target proteins with significant causal effects inferred by Mendelian randomization and with colocalization between GBMI GWAS and protein quantitative trait loci (pQTL). **B**, LocusZoom^60^ plots showing colocalization between GWAS for VTE and pQTL for F11. The fine-mapped variants are shown with their rsID.

**Figure 4.**
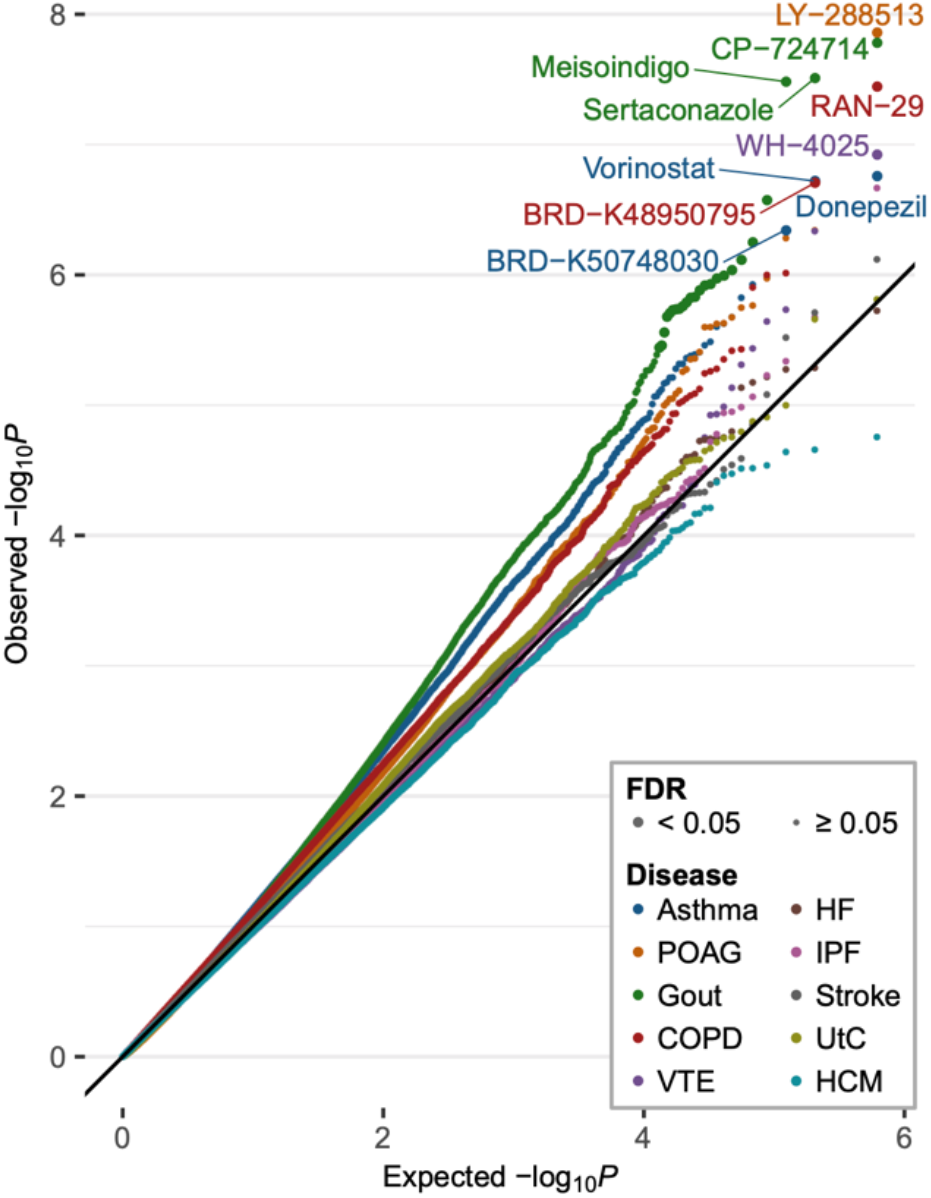
Negative correlation tests between genetically determined and compound-regulated gene expression profiles. quantile–quantile plots of the negative correlation tests between genetically determined and compound-regulated gene expression profiles. Compounds with false discovery rates (FDR) <0.05 are indicated by larger dots. The compound names are shown for at most three significant compounds, for visualization purposes.

Next, we assessed whether the inferred causal relationships were consistent with clinical implications and experimental evidence. We found literature-based support of the causal signs of the MR effect sizes for six protein–disease pairs (**Supplementary Table 3**). For example, lipoprotein(a) (LPA) for AAA and PDGFB for VTE have been reported to be disease biomarkers^35,36^. Similarly, ApoB-containing lipoproteins were associated with angiotensin II-induced AAA in a mouse model^37^. In contrast, we found that the negative sign of the MR effect size for PROC-VTE was not consistent with the knowledge that protein C, which is encoded by *PROC*, itself, is used for the treatment of VTE. The pQTL of PROC was identified as a *trans*-pQTL, which might confound the sign because PROC may not be the direct target of the pQTL effects.

We curated drugs from the four major drug databases: DrugBank^38^, Therapeutic Target Database (TTD)^39^, PharmGKB^40^, and the Open Targets Platform^41^, resulting in 83 drugs for 14 protein–disease pairs (**Supplementary Table 3**). These drugs included MAP2K inhibitors for HF, which experimentally ameliorate cardiac hypertrophy and cardiomyopathy^42,43^. Regarding the F11–VTE pair, an F11 inhibitor, Abelacimab, showed efficacy for the prevention of VTE in a phase II trial^44^. In addition, as an agonist of PLAU for asthma, the urokinase-type plasminogen activator was reported to reduce airway remodeling in a mouse model^45^.

### Negative correlation tests between genetically determined and compound-regulated gene expression

Finally, we performed an *in silico* screening of negative correlations between TWAS-based disease case–control GReX and compound-regulated gene expression profiles^16^ to identify compounds with a potentially beneficial effect for treatment of each disease. By matching the cell and tissue specificity between TWAS (based on the GTEx tissues) and compound-regulated gene expression profiles (cell lines collected in the LINCS L1000 library), we tested the negative correlation for 308,872 compound–tissue–condition pairs per disease (Methods). Because a large sample size and population-specific LD structure are required for robust *in silico* estimation of GReX by TWAS^9^, we restricted our analysis to the results of the population-specific meta-analyses of EAS and NFE. An EAS-specific meta-analysis was not performed in GBMI for three diseases, i.e., ThC, AAA, and AcApp, which were excluded. We calculated correlation coefficients in the two populations separately and subsequently combined them in a random-effect meta-analysis framework. We obtained 31 compound– disease pairs with an FDR < 0.05 (**Figure 4**).

The negative correlation tests can be applied even to compounds without known targets when the compound-induced gene expression changes are assayed. In fact, most of the prioritized compounds (14 out of 31) were understudied or had no known targets. These compounds were valuable, because they could be therapeutic drugs with different modes of action from existing drugs. Nevertheless, several prioritized compounds were well studied and had supporting evidence. A histone deacetylase (HDAC) inhibitor, vorinostat, was prioritized for asthma; concordantly, HDAC inhibition was an effective treatment in an animal model of asthma^46^.

We further identified promising compounds with marginal significance (FDR < 0.1). There were 123 unique compound–disease pairs with an FDR < 0.1, and mechanistic actions were known for 72 compounds. The supporting literature was identified for the 26 compounds. Indacaterol (a beta-2 adrenergic receptor (ADRB2) agonist) and masatinib (a proto-oncogene c-Src (SRC) inhibitor) have undergone phase III clinical trials for asthma^47,48^. There are approved drugs (acetazolamide, fluorouracil, and naproxen and indomethacin, respectively) with the same mechanistic action as cianidanol (a carbonic anhydrase inhibitor) and raltitrexed (a thymidylate synthase inhibitor) for POAG^49^, and piketoprofen and diclofenac (cyclooxygenase 1/2 inhibitors) for gout, respectively. We also prioritized several compounds currently under investigation, including phosphatidylinositol-3 kinase (PI3K) inhibitors for asthma^50^, a sodium channel inhibitor for POAG^51^, and a cyclooxygenase-2 (COX2) inhibitor for VTE^52^. We comprehensively summarized the screened drug list and their relevant evidence in **Supplementary Table 4**, which should provide genetic support to the compounds under investigation and the understudied compounds. In addition, we searched for structurally similar compounds (similarity > 0.85) for the compounds without known targets by using BindingDB^53^. We identified potential targets for six compounds (**Supplementary Table 5**), which would help investigate those compounds as therapeutic candidates.

### Combining the three approaches for in-depth drug discovery

We summarized the representative drugs and their targets, which were prioritized through the three drug discovery approaches, in **Table 1**. The overlap enrichment analysis and the endophenotype MR analysis identified 30 and 83 drugs for 13 and 14 drug targets, respectively, and 31 compounds were nominated by the negative correlation tests. Multiple components nominated drug candidates for asthma, gout, COPD, and VTE, indicating that the three components were complementarily for these diseases. We noted that these nominated diseases had a relatively large number of case sample sizes and genome-wide significant loci. The enhanced statistical power of the GWAS for a wide range of phenotypes in the framework of global biobank collaboration should contribute to the acceleration of novel drug discovery. In fact, the novel loci in GBMI^7^ led to the prioritization of *PROC* for VTE (via the overlap enrichment analysis) and PLAU for AAA (via the endophenotype MR). F11 for VTE was prioritized by both the overlap enrichment analysis and endophenotype MR, providing more robust evidence for F11 inhibitors from the genetic point of view. Conversely, no drugs were nominated for diseases with a relatively small number of loci by either the overlap enrichment analysis or negative correlation tests (e.g., IPF, UtC, and HCM). As these two components require relatively large numbers of GWAS signals in their schemes, further accumulation of samples is warranted.

**Table1.** Drug targets and representative drugs prioritized in this study.

| Disease | Overlap enrichment in disease-relevant ATC codes | Endophenotype MR | Negative correlation tests |
| --- | --- | --- | --- |
| Asthma | Ibudilast (*IL6*)<br>Omalizumab (*FCER1A/FCER1G*) | SAR164653 (CTSA)<br>KHK-2866 (HBEGF)<br>Amediplase (PLAU) | Donepezil (ACHE)<br>Vorinostat (HDAC family)<br>BRD-K50748030 (Unknown) |
| POAG | - | - | LY-288513 (Unknown) |
| Gout | Probenecid (*SLC22A6/SLC22A11*)<br>Lesinurad (*SLC22A11/SLC22A12*) | Anfibatide (GP1BA)<br>Berotralstat (KLKB1) | Mesoridazine (DRD2/HTR2A)<br>CP-724714 (Unknown)<sup>1</sup> |
| COPD | - | GMA-161 (FCGR3B) | BRD-K48950795 (Unknown)<br>RAN-29 (Unknown) |
| VTE | Alteplase (*FGA*)<br>Edoxaban (*F10*)<br>Ancrod (*PROC*)<br>Dabigatran etexilate (*F2*)<br>Abelacimab (*F11*)<br>Ecallantide (*KLKB1*)<br>Drotrecogin alfa (*F5*) | Abelacimab (F11)<br>CR-002 (PDGFB) | WH-4025 (Unknown) |
| ThC | - | Tosedostat (XPNPEP2) | (Excluded from the analysis) |
| AAA | - | MG-1102 (LPA)<br>Amediplase (PLAU) | (Excluded from the analysis) |
| HF | - | MG-1102 (LPA) | - |
| IPF | - | - | - |
| Stroke | - | - | - |
| UtC | - | Anfibatide (GP1BA) | - |
| AcApp | - | GNF-PF-5434 (CTSB) | (Excluded from the analysis) |
| HCM | - | - | - |

<sup>1</sup> Drugs without known target genes were not shown except for the drug with the lowest p-value, CP-724714. Full results can be found in **Supplementary Table 2**.
The diseases are sorted in the decreasing order of the number of the genome-wide significant loci in GBMI GWAS.

### Drug discovery nominated plausible candidate drugs and target genes for VTE

We noted that the VTE GWAS was most successful in the screening of the drug targets: it prioritized drugs corresponding to the eight drug-target genes and one compound in total (**Figure 5**). All drug targets but *PDGFB* were involved in the coagulation cascade. *PDGFB* has been reported to induce the expression of a tissue factor that triggers the coagulation cascade^54^, which underscores the strong enrichment of candidate drug targets in coagulation-related genes. Of the eight drug targets, three (*PROC, F2*, and *F10*) were targeted by the approved drugs, and two (*KLKB1* and *F11*) were targeted by drugs under clinical trials for VTE, thus supporting the validity of genomics-driven drug discovery and repositioning for VTE.

**Figure 5.**
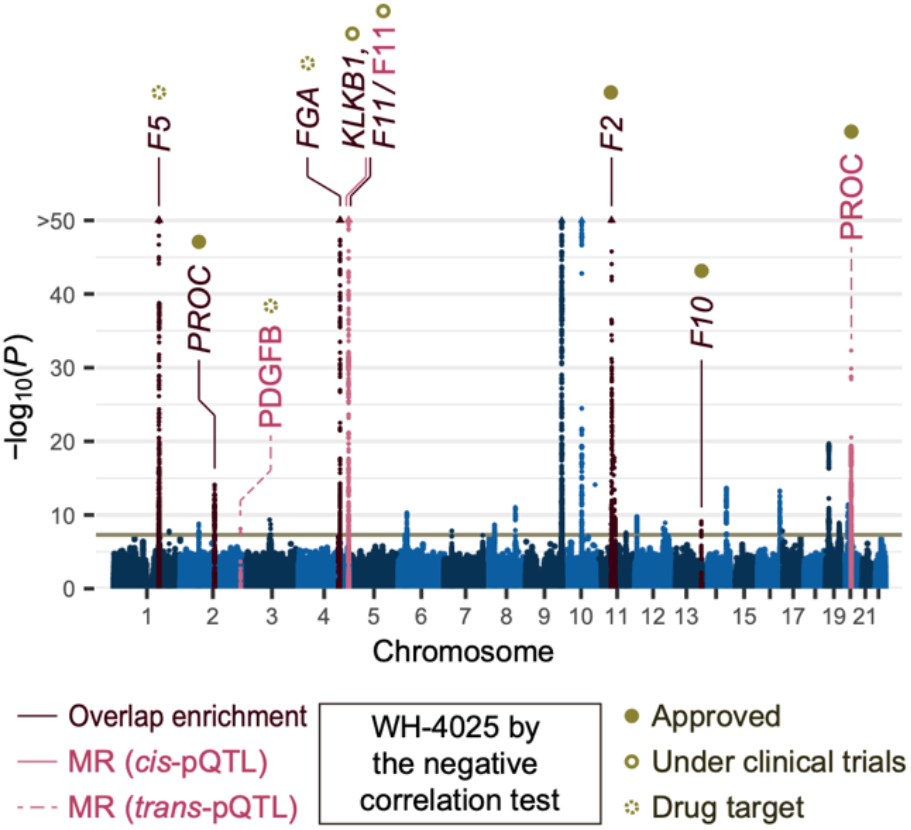
Drug discovery nominated plausible candidate drugs and target genes for VTE. Drug-target genes nominated by omnibus gene prioritization and proteins nominated by Mendelian randomization (MR) are highlighted in a Manhattan plot of GBMI GWAS for VTE. The compound with a significant negative correlation between genetically regulated and compound-regulated gene expression profiles is also shown.

## Discussion

In this study, we presented a practical framework that combined three approaches for in-depth genomics-driven drug discovery, and demonstrated its utility through application to the GBMI GWAS meta-analysis. Each approach has specific advantages. By focusing on the genes prioritized in the disease-relevant medication codes, we obtained a list of candidate drugs, most of which were indicated for the diseases. Endophenotype MR and subsequent quality controls estimated the causality of proteins regarding diseases, which could not be inferred by gene prioritization. Finally, the negative correlation tests of gene expression profiles were able to nominate compounds with or without known target genes. Our study demonstrated the importance of combining the three components for the thorough evaluation of candidate drugs.

Multi-ethnic GWAS meta-analyses incorporated populations with diverse genetic backgrounds and architectures. Matching of genetic ancestry is important to follow-up functional interpretation of the GWAS results with omics information, including drug discovery. To address the difference in the LD structure between populations, we used the GWAS summary statistics from the population-specific meta-analysis for endophenotype MR and the negative correlation tests, whereas we used those from the all-population meta-analyses for gene prioritization. The differences in ancestry-matching strategies among components depended on the current availability of the corresponding omics resource requested for each analysis. Further accumulation of public resources with diverse ancestry should be expected^25,55^.

A large number of GWAS loci were required for genomics-driven drug discovery, especially for the overlap enrichment analysis and the negative correlation tests. Global biobank collaborations, such as GBMI, have the potential to improve the power of genetic association studies to detect novel GWAS signals by incorporating diverse populations with large sample sizes, and will facilitate genomics-driven drug discovery. Of note, our framework successfully nominated drug candidates and their target genes, particularly for VTE, which had the fifth-largest number of GWAS loci among the 13 diseases. There may be additional factors other than the number of GWAS loci that are important for the success of drug discovery in VTE. Among the traits with a large number of loci, VTE showed a relatively modest polygenicity^7^, which might be beneficial for pinpointing the disease-relevant genes. The genes implicated in the VTE GWAS were mainly centered at the coagulation cascade^56^. The drugs targeting coagulation factors have been under active development^57^, and these drugs can be promising candidates immediately repositioned to coagulation disorders other than the disease for which the drugs were originally developed. The further evaluation of the suitability of drug discovery for a broader range of phenotypes, including cancer, autoimmune diseases, and coagulation disorders would be an interesting direction for future research.

There are several potential limitations for each drug discovery component. The overlap enrichment analysis required the existence of the drugs approved for the disease; therefore, its application to diseases without approved drugs would be challenging. Endophenotype MR could be applied to proteins targeted by pQTL studies; however, the number of proteins in the pQTL studies is currently limited because of the technological difficulty of proteomics^58^. Regarding the negative correlation tests, the LINCS L1000 compound library does not contain gene expression profiles for all pairs of compounds and cell lines^21^. Therefore, we used all tissue types in GTEx, regardless of disease relevance. In addition, the methodological limitations of TWAS^9,59^ might affect drug discovery using GReX.

In conclusion, our drug discovery framework practically afforded the *in silico* screening of abundant drugs and targets with supporting evidence. It enables the routine to conduct the post-GWAS genomics-driven drug discovery in the era of cross-population GWAS meta-analysis, which would further facilitate the translation of GWAS findings to therapeutic targets.

## Methods

### GBMI GWAS meta-analysis

GBMI GWAS is a meta-analysis of 18 biobanks incorporating up to 1.8 million participants with diverse ancestries (341,000 EAS; 31,000 Central and South Asians; 33,000 Africans; 18,000 Admixed Americans; 1,600 Middle Easterners; 156,000 Finns; and 1,220,000 NFE)^7^. We used the GBMI GWAS of 13 common diseases: asthma, POAG, gout, COPD, VTE, ThC, AAA, HF, IPF, stroke, UtC, AcApp, and HCM. Although a GBMI GWAS was also conducted for appendectomy, we excluded this trait because it was a procedure endpoint rather than a disease.

### Gene prioritization

We used four tools for gene prioritization from GWAS summary statistics, i.e., MAGMA, DEPICT, Pi, and PoPS. We used the default settings, unless otherwise stated. Variants with *P*-values < 1.0 × 10^−5^ were used as input for DEPICT. We followed the original paper for the setup of Pi^5^. Specifically, we used the lead variants with *P* < 5.0×10^−8^ as input, eQTL in the peripheral blood and immune cells, chromatin interaction in immune cells, topologically associating domain boundary in the GM12878 cell line, and the STRING^61^ protein–protein interaction network, with a high confidence score. PoPS was originally developed to pinpoint one responsible gene per locus, and chooses the gene with the highest score at the locus of interest^24^. Here, we used top-ranked genes, rather than pinpointed genes, to incorporate multiple genes per locus for drug discovery. We used the summary statistics of the all-population GWAS meta-analyses as input and the European subset of the 1000 Genomes Project (Phase 3) as a reference, given that more than half of the GBMI samples were NFE^7^. We prioritized genes with conventional thresholds, i.e., FDR < 0.05 for MAGMA, and top 5% of the genes in the descending order of gene scores for Pi and PoPS. We used an FDR < 0.2 for DEPICT, as DEPICT calculates *P*-values only for the genes in the pre-featured target loci, by default. When we examined sequentially changed thresholds as a sensitivity analysis, we used FDR thresholds of 0.001, 0.005, 0.01, 0.05, 0.1, 0.2, and 0.5 for MAGMA and DEPICT. Pi and PoPS calculate the gene score instead of FDR. We used the top 1%, 5%, and 10% of the genes with the highest gene scores. In addition, a Pi score > 1.5, 2.0, 2.5, and 3.0 for Pi, and a PoPS score > 0, 0.5, 1.0, 1.5, and 2 for PoPS were also examined.

### Enrichment analysis of drug indication categories

We used the list of prioritized genes as input to perform a series of Fisher’s exact tests for ATC or ICD-10 codes, to test the enrichment of drug-target genes in particular codes. We used the drug-target database provided by GREP^14^, which was constructed by curating two major drug databases, Drug Bank and TTD. The ICD-10 codes were summarized into the 21 large categories, as shown in **Supplementary Figure 2**. Because ICD-10 is a disease-classification system, we simply defined the relevant ICD-10 category as the category that contained the disease. Conversely, ATC is a drug-classification system, and the approved drugs can belong to multiple ATC codes. Therefore, we defined the disease-relevant ATC code as the ATC code with the largest number of approved drugs for the disease-relevant ICD-10 category. We defined the disease-irrelevant ATC codes as the ATC codes without any approved drugs for the disease-relevant ICD-10 category. Regarding ATC, we limited the enrichment analyses to the diseases for which there were more than four approved drugs in the disease-relevant ATC codes. As a result, four diseases (i.e., AAA, IPF, AcApp, and HCM) were excluded.

### Mendelian randomization of the pQTL signals

We used Wald ratio tests for MR analyses. The lead variants of the five protein QTL studies^27– 31^ were evaluated as described previously^18^. We used the lead variants of drug-target proteins classified as tier 1 variants as instrumental variables. The tier 1 variants were defined as the variants that were associated with no more than five proteins and did not show heterogeneity in the five studies. When the lead variant of the pQTL was missing in the GBMI GWAS summary statistics, we used a proxy variant with the largest *R*^*2*^, if the *R*^*2*^ was larger than 0.8. We checked the directionality of causal relationships using Steiger filtering^62^. MR analyzes were performed using the “TwoSampleMR” R package^63^.

### Colocalization analysis

To test colocalization in the presence of multiple causal variants, we applied coloc to the signals decomposed by SuSiE^32^ for each locus, including the variants located within ±500 kb of the lead variant (coloc + SuSiE). If SuSiE^34^ did not converge in 100 iterations for either pQTL or GBMI GWAS, we instead used coloc^33^. Coloc + SuSiE and coloc were performed using their default parameters. The summary statistics were not publicly available for two pQTL studies^30,31^; therefore, we compared *R*^*2*^ between the lead variants of the pQTL and the GBMI GWAS for those studies. We considered that the signals of pQTL and GBMI GWAS were colocalized if the maximum posterior probability of colocalization (i.e., PP.H4 for coloc and coloc + SuSiE) was larger than 0.8, or the *R*^*2*^ between the lead variants was larger than 0.8.

### Negative correlation tests between genetically determined and compound-regulated gene expression

We utilized Trans-Phar^16^ for the negative correlation tests. Trans-Phar internally used FOCUS^64^ to infer disease case–control GReX for 44 GTEx v7 tissues based on the GWAS summary statistics. Compound-regulated gene expression profiles were obtained from the LINCS L1000 library. We calculated Spearman’s rho between GReX and compound-regulated gene expression profiles using the GReX inferred from the NFE-specific and EAS-specific GWAS meta-analyses, separately. The correlation coefficients were combined by a random-effect model using the R package “metacor.”

### Filtering GWAS summary statistics based on effective sample sizes

Because of the diverse ancestry in the GWASs for meta-analysis, there was remarkable heterogeneity in the effective sample sizes across the genome-wide variants of the GBMI results of each phenotype. This heterogeneity affected the performance of downstream analyses, including those of polygenic risk score^65^. Therefore, we excluded variants with effective sample sizes <50% of the maximum effective sample size from the GWAS summary statistics of each phenotype.

## Data Availability

All data used in the present study are publicly available

## Data and code availability

The GBMI GWAS summary statistics are publicly available at https://www.globalbiobankmeta.org/resources. The genomics-driven drug discovery analysis was conducted using the following publicly available tools: MAGMA (https://ctg.cncr.nl/software/magma), DEPICT (https://data.broadinstitute.org/mpg/depict/), PoPS (https://github.com/FinucaneLab/pops), GREP (https://github.com/saorisakaue/GREP), Trans-Phar (https://github.com/konumat/Trans-Phar), LocusZoom (http://locuszoom.org/), and the Pi, TwoSampleMR, coloc, susieR, and metacor R packages.

## Author contributions

S.N., T.K. and Y.O. designed the study, and wrote the manuscript. S.N., T.K., K.H.W., and W.Z. conducted data analysis. Y.O. supervised the study.

## Declare of interests

The authors declare no competing interests.

## References

1. Hay, M., Thomas, D. W., Craighead, J. L., Economides, C. & Rosenthal, J. Clinical development success rates for investigational drugs. Nat. Biotechnol. 32, 40–51 (2014).

2. Nelson, M. R. et al. The support of human genetic evidence for approved drug indications. Nat. Genet. 47, 856–860 (2015).

3. King, E. A., Davis, J. W. & Degner, J. F. Are drug targets with genetic support twice as likely to be approved? Revised estimates of the impact of genetic support for drug mechanisms on the probability of drug approval. PLOS Genet. 15, e1008489 (2019).

4. Sabatine, M. S. et al. Evolocumab and Clinical Outcomes in Patients with Cardiovascular Disease. N. Engl. J. Med. 376, 1713–1722 (2017).

5. Fang, H. et al. A genetics-led approach defines the drug target landscape of 30 immune-related traits. Nat. Genet. 51, 1082–1091 (2019).

6. Reay, W. R. & Cairns, M. J. Advancing the use of genome-wide association studies for drug repurposing. Nat. Rev. Genet. 22, 658–671 (2021).

7. Zhou, W. et al. Global Biobank Meta-analysis Initiative: power genetic discovery for human diseases with > 2.6 million samples across diverse ancestries. medRxiv (2021) doi:10.1101/2021.11.19.21266436.

8. Sakaue, S. et al. A cross-population atlas of genetic associations for 220 human phenotypes. Nat. Genet. 53, 1415–1424 (2021).

9. Wainberg, M. et al. Opportunities and challenges for transcriptome-wide association studies. Nat. Genet. 51, 592–599 (2019).

10. Kuchenbaecker, K. et al. The transferability of lipid loci across African, Asian and European cohorts. Nat. Commun. 10, 4330 (2019).

11. Shi, H. et al. Population-specific causal disease effect sizes in functionally important regions impacted by selection. Nat. Commun. 12, 1098 (2021).

12. Okada, Y. et al. Genetics of rheumatoid arthritis contributes to biology and drug discovery. Nature 506, 376–381 (2013).

13. Malik, R. et al. Multiancestry genome-wide association study of 520,000 subjects identifies 32 loci associated with stroke and stroke subtypes. Nat. Genet. 50, 524–537 (2018).

14. Sakaue, S. & Okada, Y. GREP: Genome for REPositioning drugs. Bioinformatics 35, 3821–3823 (2019).

15. Zhou, S. et al. A Neanderthal OAS1 isoform Protects Against COVID-19 Susceptibility and Severity: Results from Mendelian Randomization and Case-Control Studies. medRxiv (2020) doi:10.1101/2020.10.13.20212092.

16. Konuma, T., Ogawa, K. & Okada, Y. Integration of genetically regulated gene expression and pharmacological library provides therapeutic drug candidates. Hum. Mol. Genet. 30, 294–304 (2021).

17. Sonehara, K. & Okada, Y. Genomics-driven drug discovery based on disease-susceptibility genes. doi:10.1186/s41232-021-00158-7.

18. Zheng, J. et al. Phenome-wide Mendelian randomization mapping the influence of the plasma proteome on complex diseases. Nat. Genet. 52, 1122–1131 (2020).

19. So, H.-C. et al. Analysis of genome-wide association data highlights candidates for drug repositioning in psychiatry. Nat. Neurosci. 20, 1342–1349 (2017).

20. Aguet, F. et al. Genetic effects on gene expression across human tissues. Nature 550, 204–213 (2017).

21. Subramanian, A. et al. A Next Generation Connectivity Map: L1000 Platform and the First 1,000,000 Profiles. Cell 171, 1437-1452.e17 (2017).

22. Leeuw, C. A. de, Mooij, J. M., Heskes, T. & Posthuma, D. MAGMA: Generalized Gene-Set Analysis of GWAS Data. PLOS Comput. Biol. 11, e1004219 (2015).

23. Pers, T. H. et al. Biological interpretation of genome-wide association studies using predicted gene functions. Nat. Commun. 6, 5890 (2015).

24. Weeks, E. M. et al. Leveraging polygenic enrichments of gene features to predict genes underlying complex traits and diseases. medRxiv 23, (2020).

25. Zhao, H. et al. Proteome-wide Mendelian randomization in global biobank meta-analysis reveals trans-ancestry drug targets for common diseases. medRxiv (2021).

26. Võsa, U. et al. Large-scale cis-and trans-eQTL analyses identify thousands of genetic loci and polygenic scores that regulate blood gene expression. Nat. Genet. 2021 539 53, 1300–1310 (2021).

27. Sun, B. B. et al. Genomic atlas of the human plasma proteome. Nature 558, 73–79 (2018).

28. Suhre, K. et al. Connecting genetic risk to disease end points through the human blood plasma proteome. Nat. Commun. 8, 14357 (2017).

29. Folkersen, L. et al. Mapping of 79 loci for 83 plasma protein biomarkers in cardiovascular disease. PLOS Genet. 13, e1006706 (2017).

30. Emilsson, V. et al. Co-regulatory networks of human serum proteins link genetics to disease. Science (80-.). 361, 769–773 (2018).

31. Yao, C. et al. Genome-wide mapping of plasma protein QTLs identifies putatively causal genes and pathways for cardiovascular disease. Nat. Commun. 9, 3268 (2018).

32. Wallace, C. A more accurate method for colocalisation analysis allowing for multiple causal variants. PLOS Genet. 17, e1009440 (2021).

33. Giambartolomei, C. et al. Bayesian Test for Colocalisation between Pairs of Genetic Association Studies Using Summary Statistics. PLOS Genet. 10, e1004383 (2014).

34. Wang, G., Sarkar, A., Carbonetto, P. & Stephens, M. A simple new approach to variable selection in regression, with application to genetic fine mapping. J. R. Stat. Soc. Ser. B (Statistical Methodol. 82, 1273–1300 (2020).

35. Kotani, K. et al. Lipoprotein(a) Levels in Patients With Abdominal Aortic Aneurysm. Angiology 68, 99–108 (2017).

36. Bruzelius, M. et al. PDGFB, a new candidate plasma biomarker for venous thromboembolism: results from the VEREMA affinity proteomics study. Blood 128, e59–e66 (2016).

37. Liu, J. et al. Associations of ApoAI and ApoB–Containing Lipoproteins With AngII– Induced Abdominal Aortic Aneurysms in Mice. Arterioscler. Thromb. Vasc. Biol. 35, 1826–1834 (2015).

38. Wishart, D. S. et al. DrugBank 5.0: a major update to the DrugBank database for 2018. Nucleic Acids Res. 46, D1074–D1082 (2018).

39. Chen, X., Ji, Z. L. & Chen, Y. Z. TTD: Therapeutic Target Database. Nucleic Acids Res. 30, 412–415 (2002).

40. Whirl-Carrillo, M. et al. An Evidence-Based Framework for Evaluating Pharmacogenomics Knowledge for Personalized Medicine. Clin. Pharmacol. Ther. 110, 563–572 (2021).

41. Ochoa, D. et al. Open Targets Platform: supporting systematic drug–target identification and prioritisation. Nucleic Acids Res. 49, D1302–D1310 (2021).

42. Sala, V. et al. Cardiac concentric hypertrophy promoted by activated Met receptor is mitigated in vivo by inhibition of Erk1,2 signalling with Pimasertib. J. Mol. Cell. Cardiol. 93, 84–97 (2016).

43. A, M., W, W., F, S., S, H. & HJ, W. Mitogen-activated protein kinase kinase 1/2 inhibition and angiotensin II converting inhibition in mice with cardiomyopathy caused by lamin A/C gene mutation. Biochem. Biophys. Res. Commun. 452, 958–961 (2014).

44. Verhamme, P. et al. Abelacimab for Prevention of Venous Thromboembolism. N. Engl. J. Med. 385, 609–617 (2021).

45. Kuramoto, E. et al. Inhalation of urokinase-type plasminogen activator reduces airway remodeling in a murine asthma model. Am. J. Physiol. Cell. Mol. Physiol. 296, L337–L346 (2009).

46. Ren, Y. et al. Therapeutic effects of histone deacetylase inhibitors in a murine asthma model. Inflamm. Res. 65, 995–1008 (2016).

47. van Zyl-Smit, R. N. et al. Once-daily mometasone plus indacaterol versus mometasone or twice-daily fluticasone plus salmeterol in patients with inadequately controlled asthma (PALLADIUM): a randomised, double-blind, triple-dummy, controlled phase 3 study. Lancet Respir. Med. 8, 987–999 (2020).

48. Chanez, P. et al. Masitinib Significantly Decreases the Rate of Asthma Exacerbations in Patients with Severe Asthma Uncontrolled by Oral Corticosteroids: A Phase 3 Multicenter Study. in B93. LATE BREAKING CLINICAL TRIALS IN AIRWAY DISEASES A4210–A4210 (American Thoracic Society, 2020). doi:10.1164/ajrccm-conference.2020.201.1_MeetingAbstracts.A4210.

49. Lo Faro, V. et al. Global Biobank Meta-Analysis Initiative: A genome-wide association meta-analysis identifies novel primary open-angle glaucoma loci and shared biology with vascular and cell proliferation mechanisms. medRxiv (2021).

50. Yoo, E. J., Ojiaku, C. A., Sunder, K. & Panettieri, R. A. Phosphoinositide 3-Kinase in Asthma: Novel Roles and Therapeutic Approaches. Am. J. Respir. Cell Mol. Biol. 56, 700–707 (2017).

51. Hains, B. C. & Waxman, S. G. Neuroprotection by Sodium Channel Blockade with Phenytoin in an Experimental Model of Glaucoma. Investig. Opthalmology Vis. Sci. 46, 4164 (2005).

52. Anderson, D. R. et al. Aspirin or Rivaroxaban for VTE Prophylaxis after Hip or Knee Arthroplasty. N. Engl. J. Med. 378, 699–707 (2018).

53. Gilson, M. K. et al. BindingDB in 2015: A public database for medicinal chemistry, computational chemistry and systems pharmacology. Nucleic Acids Res. 44, D1045–D1053 (2016).

54. Gebhard, C. et al. PDGF-CC induces tissue factor expression: role of PDGF receptor α/β. Basic Res. Cardiol. 105, 349–356 (2010).

55. Martin, A. R. et al. Clinical use of current polygenic risk scores may exacerbate health disparities. Nat. Genet. 51, 584–591 (2019).

56. Wolford, B. N. et al. Multi-ancestry GWAS for venous thromboembolism identifies novel loci followed by experimental validation. medRxiv (2021).

57. Grover, S. P. & Mackman, N. Intrinsic Pathway of Coagulation and Thrombosis. Arterioscler. Thromb. Vasc. Biol. 39, 331–338 (2019).

58. Pietzner, M. et al. Mapping the proteo-genomic convergence of human diseases. Science (80-.). 374, eabj1541 (2021).

59. Leeuw, C. de, Werme, J., Savage, J., Peyrot, W. & Posthuma, D. Reconsidering the validity of transcriptome-wide association studies. bioRxiv (2021) doi:10.1101/2021.08.15.456414.

60. Pruim, R. J. et al. LocusZoom: regional visualization of genome-wide association scan results. Bioinformatics 26, 2336 (2010).

61. Szklarczyk, D. et al. The STRING database in 2011: functional interaction networks of proteins, globally integrated and scored. Nucleic Acids Res. 39, (2011).

62. Hemani, G., Tilling, K. & Smith, G. D. Orienting the causal relationship between imprecisely measured traits using GWAS summary data. PLOS Genet. 13, e1007081 (2017).

63. Hemani, G. et al. The MR-base platform supports systematic causal inference across the human phenome. Elife 7, 1–29 (2018).

64. Mancuso, N. et al. Probabilistic fine-mapping of transcriptome-wide association studies. Nat. Genet. 51, 675–682 (2019).

65. Wang, Y. et al. Global biobank analyses provide lessons for computing polygenic risk scores across diverse cohorts. medRxiv (2021) doi:10.1101/2021.11.18.21266545.

