## Supplementary Information for "A practical guideline of genomics-driven drug discovery in the era of global biobank meta-analysis"

S Namba and T Konuma et al.

Corresponding to Yukinori Okada

### Supplementary Figures

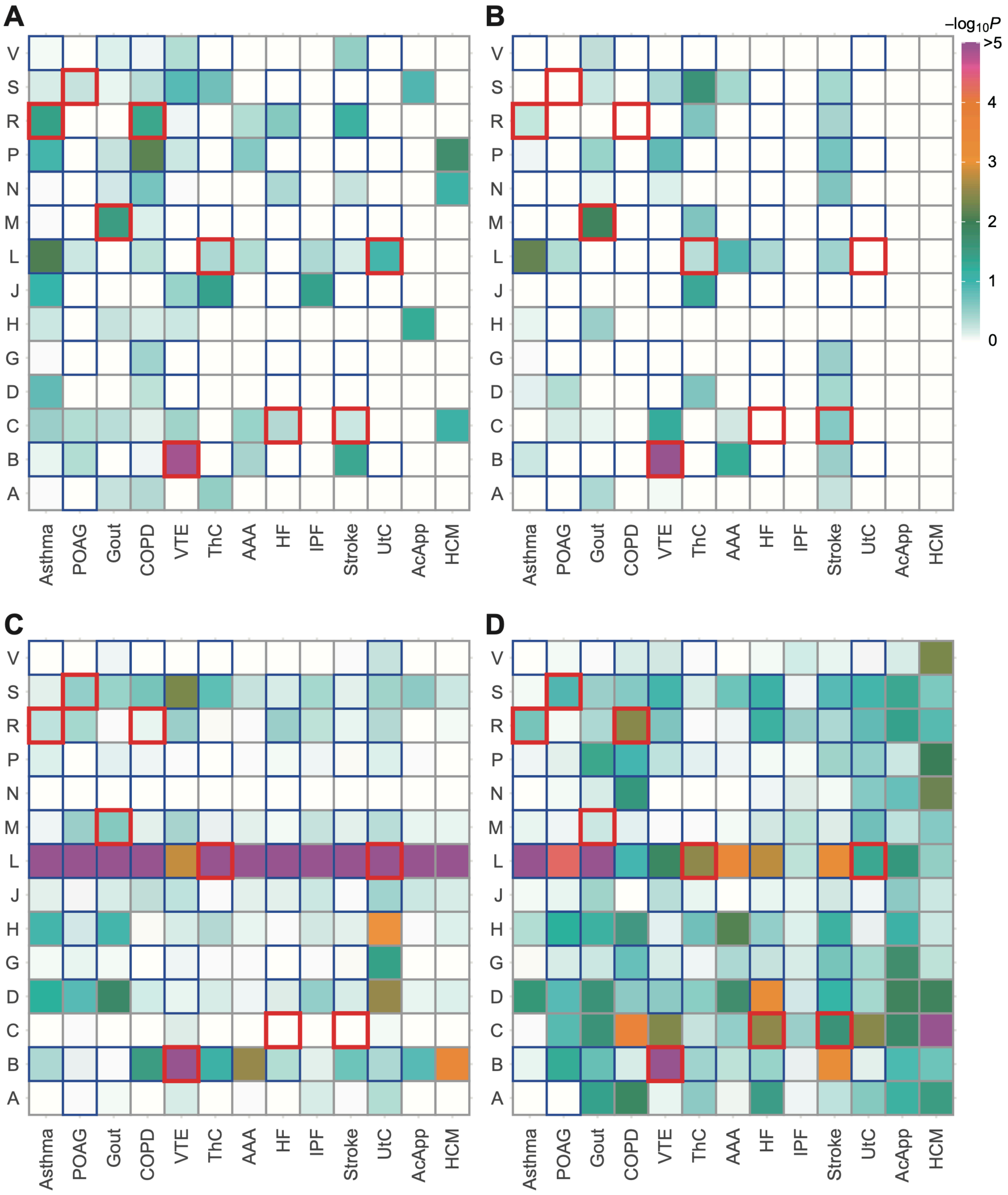

**Supplementary Figure 1, Overlap enrichment of disease risk genes with drug-target**

**genes in the ATC codes.**

Disease risk genes were prioritized by MAGMA (**A**), DEPICT (**B**), Pi (**C**), and PoPS (**D**). The disease-relevant codes and disease-irrelevant codes are circled in red and blue, respectively. The diseases are sorted in the descending order of the number of the genome-wide significant loci.

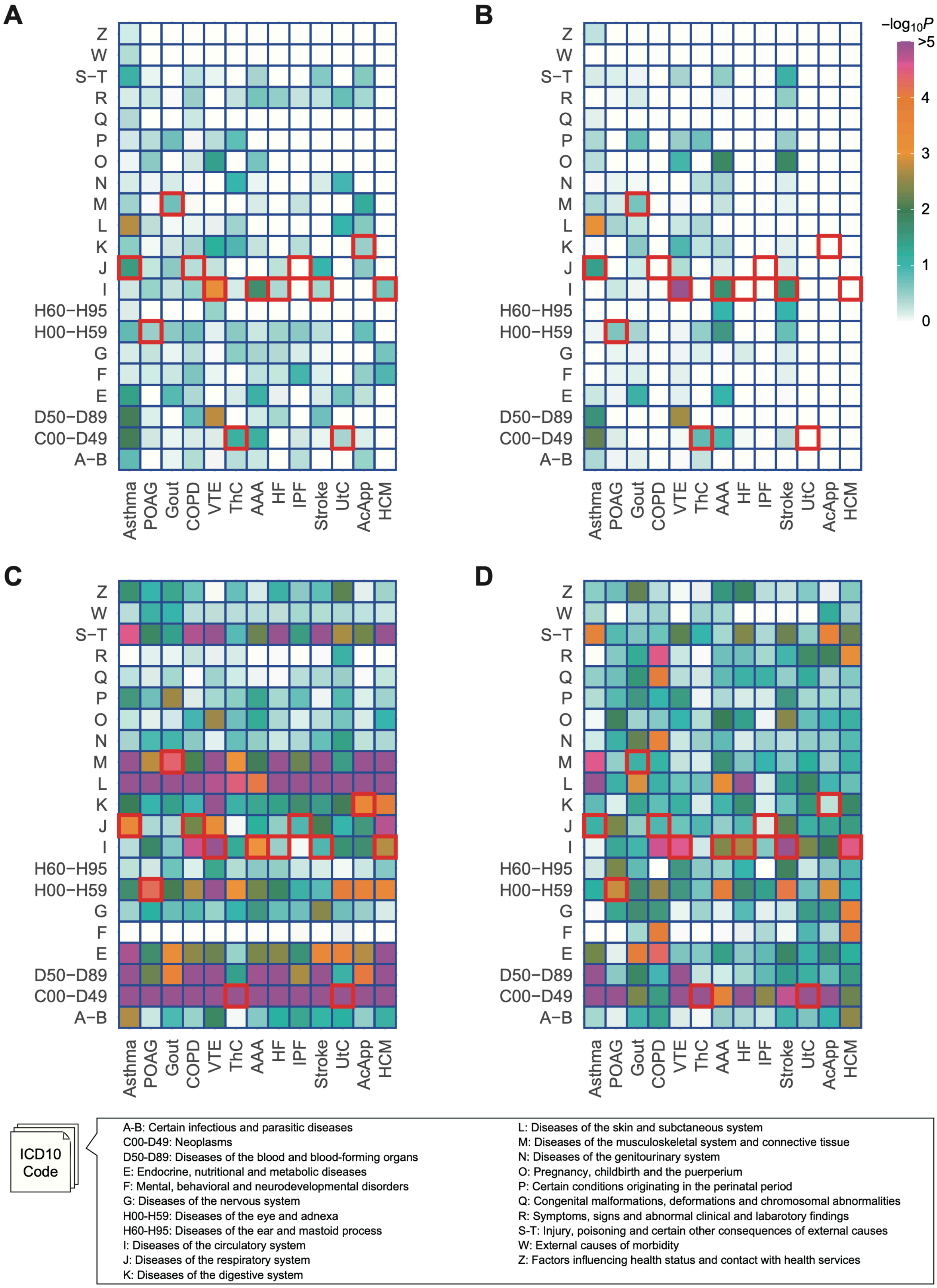
A
B
C
D
Z
W
S−T
R
Q
P
O
N
M
L
K
J
I
H60−H95
H00−H59
G
F
E
D50−D89
C00−D49
A−B
Asthma
POAG
Gout
COPD
VTE
ThC
AAA
HF
IPF
Stroke
UtC
AcApp
HCM
−log10P
>5
4
3
2
1
0
ICD10 Code
A-B: Certain infectious and parasitic diseases
C00-D49: Neoplasms
D50-D89: Diseases of the blood and blood-forming organs
E: Endocrine, nutritional and metabolic diseases
F: Mental, behavioral and neurodevelopmental disorders
G: Diseases of the nervous system
H00-H59: Diseases of the eye and adnexa
H60-H95: Diseases of the ear and mastoid process
I: Diseases of the circulatory system
J: Diseases of the respiratory system
K: Diseases of the digestive system
L: Diseases of the skin and subtaneous system
M: Diseases of the musculoskeletal system and connective tissue
N: Diseases of the genitourinary system
O: Pregnancy, childbirth and the puerperium
P: Certain conditions originating in the perinatal period
Q: Congenital malformations, deformations and chromosomal abnormalities
R: Symptoms, signs and abnormal clinical and labarotory findings
S-T: Injury, poisoning and certain other consequences of external causes
W: External causes of morbidity
Z: Factors influencing health status and contact with health services

**Supplementary Figure 2, Overlap enrichment of disease risk genes with drug-target genes in the ICD-10 codes.**

Disease risk genes were prioritized by MAGMA (**A**), DEPICT (**B**), Pi (**C**), and PoPS (**D**). The disease-relevant codes and disease-irrelevant codes are circled in red and blue, respectively. A list of ICD-10 large classification categories (i.e., chapters) are shown in the bottom.

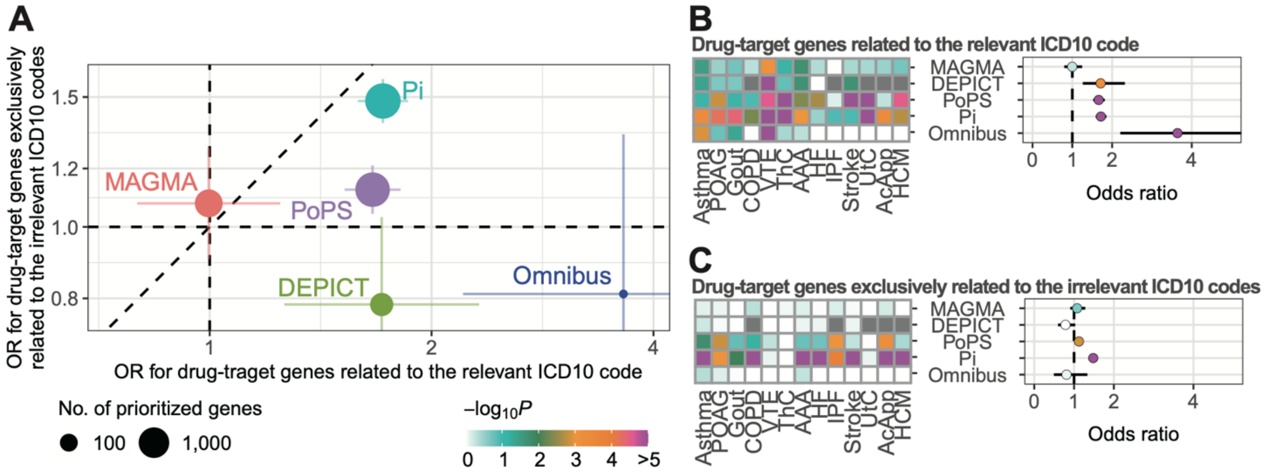

**Supplementary Figure 3, Enrichment of prioritized drug-target genes in the ICD-10 codes.**

**A**, Overall enrichments of drug-target genes nominated by four gene prioritization tools and their omnibus results in ICD-10 codes. Error bars represent 95% confidence intervals. **B and C**, enrichments of the prioritized drug-target genes in the disease-relevant ICD codes (**B**) and the disease-irrelevant ATC codes (**C**).

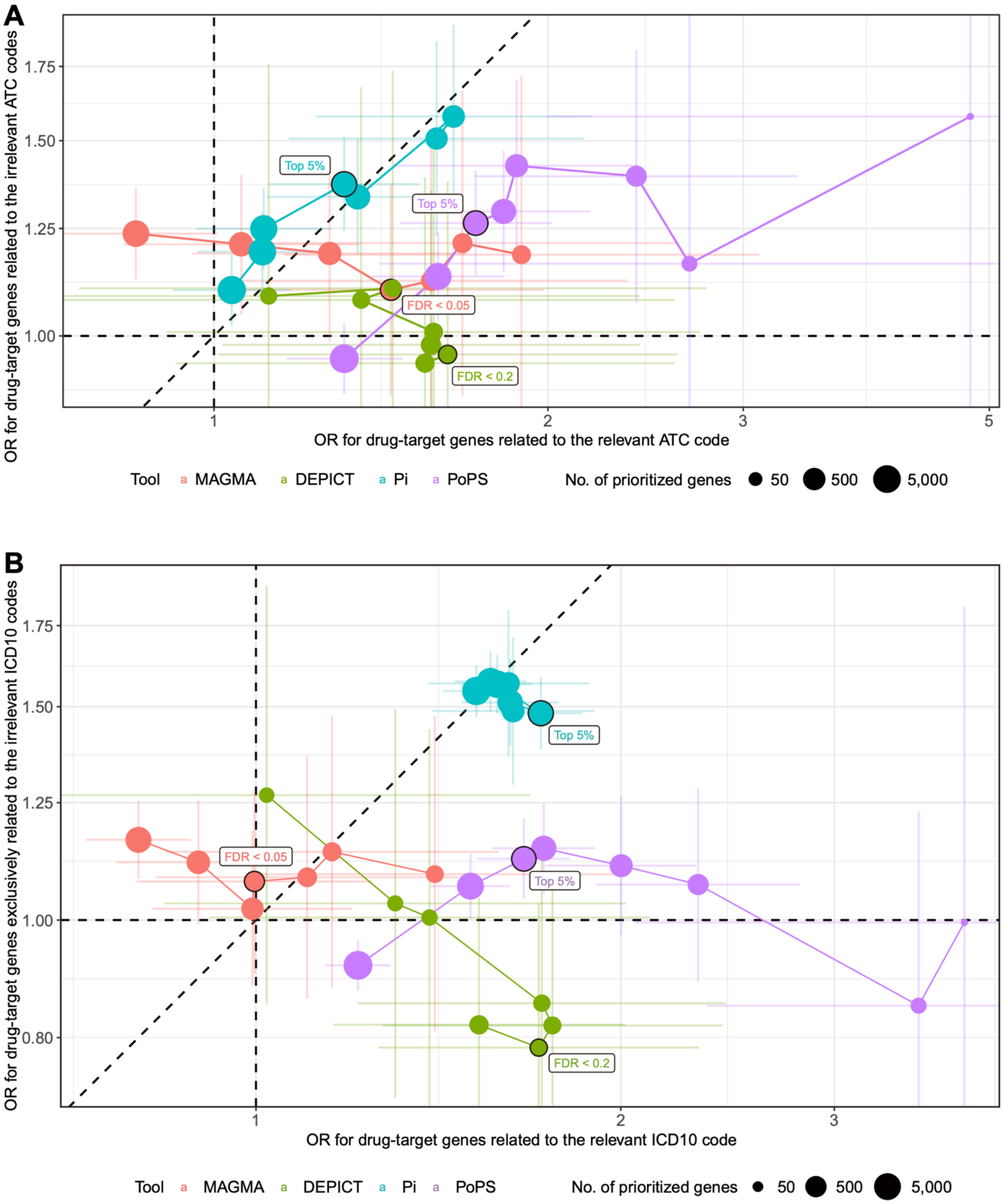

**Supplementary Figure 4, Enrichment of prioritized drug-target genes using different thresholds for gene prioritization.**

Overall enrichments of drug-target genes in the disease-relevant and disease-irrelevant medication categories with different thresholds for gene prioritization. ATC codes and ICD-10 codes were used as the medication categories for (**A**) and (**B**), respectively. The points are connected in the order of the number of prioritized genes. The thresholds used in the main analyses are labeled.
